# An Evaluation of the MedSnap Medication Authentication System

**DOI:** 10.1101/2020.05.06.20093427

**Authors:** Patrick Hymel, Daisy Y. Wong, Ning Zheng, Stephen E. Brossette

## Abstract

Falsified and substandard (SF) medications are a large and growing global public health threat affecting developed and developing markets. The need for scalable, easy-to-use, accurate methods to detect SF medications continues to grow. We conducted a proof-of-principle laboratory and field sample evaluation of MedSnap in Vientiane, Lao PDR. Over three days, MedSnap models were created from trusted and authentic medications and tested against samples of authentic/trusted or falsified artesunate, artemether-lumefantrine, azithromycin, and ciprofloxacin. Across 48 tests of authentic/trusted and falsified samples, MedSnap was 100% sensitive and specific. On the final day of the study, a convenience sample of ciprofloxacin and azithromycin was collected from local pharmacies and tested. Each field sample matched an authentic/trusted model. The speed and ease of data collection, reference model building, and sample testing suggest scalable use cases for the system.

## Introduction

Falsified and substandard (SF) medications are a growing global public health threat, harming patients and promoting antimicrobial resistance^1^. There has been a recent deluge of SF medicines and medical products for management of COVID-19 infection, and this issue is likely to worsen^2^. Many portable technologies exist to protect the supply chain as medications are manufactured and transported for distribution^3^. However, surveillance and testing at the end of the supply chain remains difficult^1^. Clinicians, public health officials, pharmacists, and law enforcement agents need accurate, scalable, easy-to-use, and portable screening technologies.

**Figure 1.**
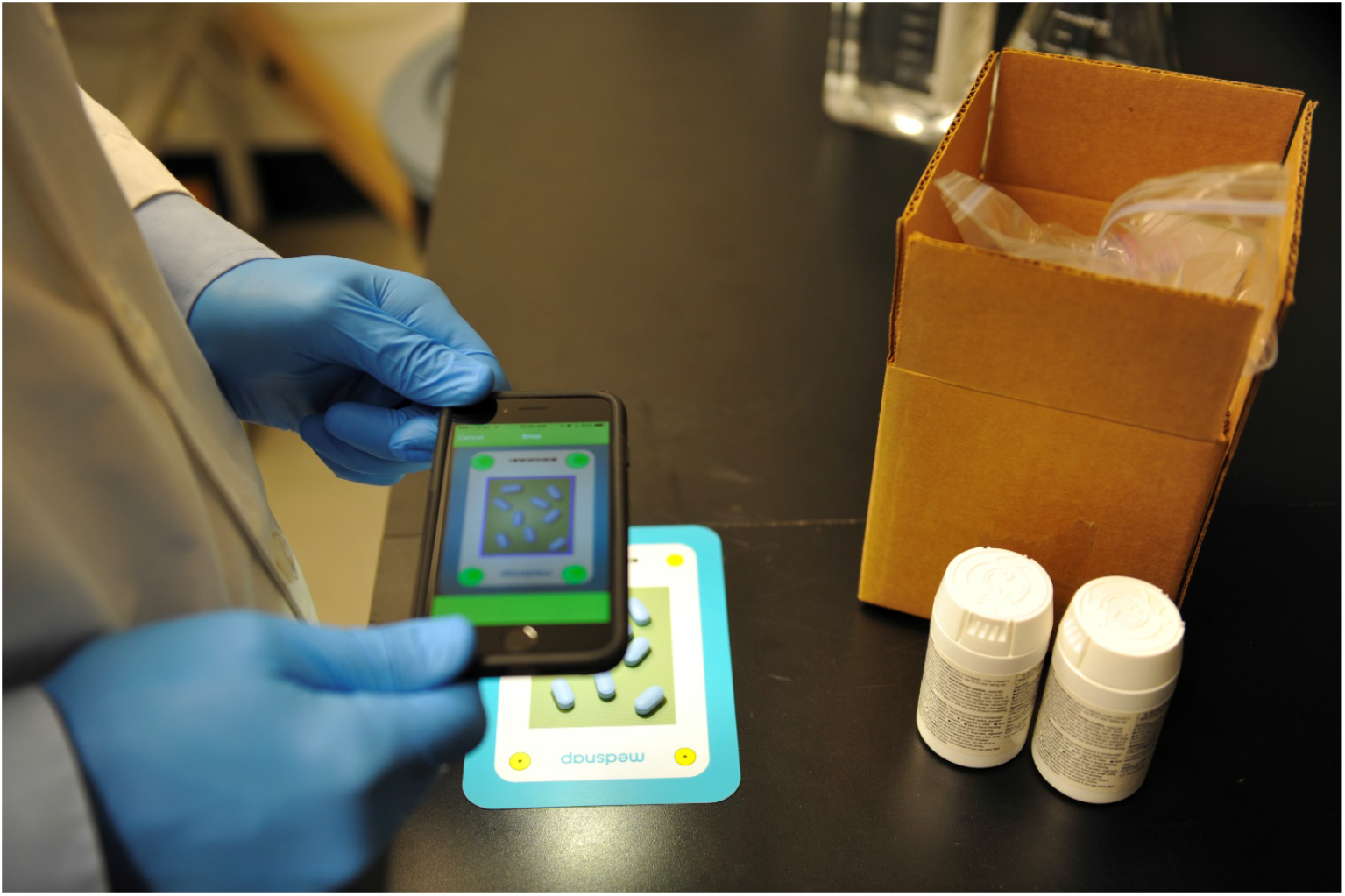
Use of the MedSnap VR application to test a sample of tablets to determine authenticity. The vinyl Snap Surface is used as a background for the sample.

MedSnap (Indicator Sciences, LLC, Seattle, WA, USA) is a smartphone-based system that uses computer vision to evaluate the authenticity of solid oral medications. MedSnap is comprised of two smartphone applications: MedSnap VR and MedSnap DA. MedSnap DA is used to acquire images of authentic samples of tablets and capsules and is used only by MedSnap or project-specific designated technical personnel. Images are uploaded to cloud-based servers where statistical models of 25 visual features are built. MedSnap VR then uses those models to evaluate the precise appearance of tablets and capsules and returns results on whether or not the tested sample is visually consistent with known authentic products.

Both applications are secured by user credentials and securely transmit data when connected to cellular or Wi-Fi networks. MedSnap VR only requires network connectivity for periodic application updates and to send results to secure cloud servers. It does not require connectivity to test samples. MedSnap VR can be automatically disabled after extended off-network use and can be remotely disabled if inappropriate use is suspected.

The applications acquire images in reasonable diffuse lighting conditions (no direct sunlight or harsh shadows), and both automatically check ambient lighting before each snap to make sure lighting is within acceptable limits.

**Figure 2.**
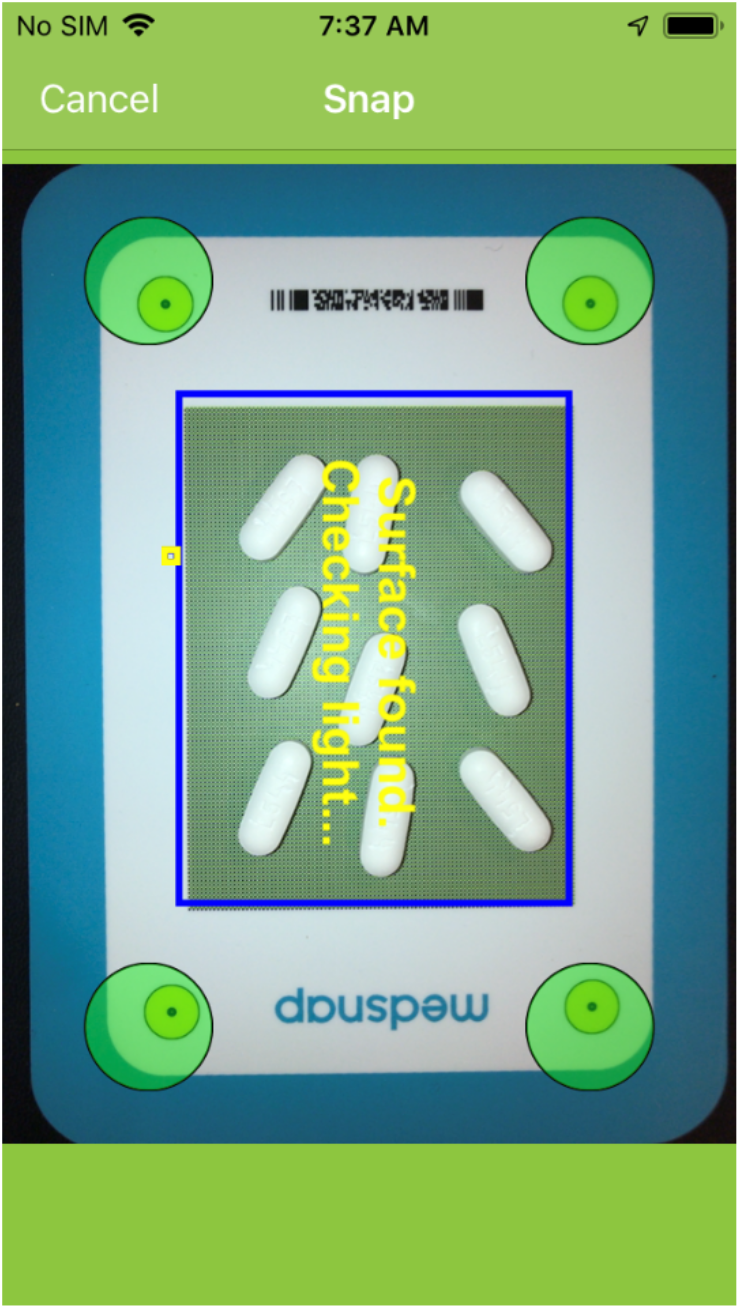
MedSnap DA and VR guide the user to properly align the camera with the Snap Surface then automatically take an image.

For testing using MedSnap VR, the user selects a medication then holds the smartphone above a sample of up to 9 tablets or capsules arranged on a durable vinyl background called a Snap Surface. Once positioned, the application automatically takes an image with flash exposure. When 9 tablet images are collected for each side, the application computes 25 features of visual appearance per side, and compares them to statistical models of known authentic medications. The VR and DA applications measure sizes to within +/− 0.1mm, construct detailed imprint composites, and differentiate 250,000+ shades of color.

## Methods

We conducted a proof-of-principle laboratory and field sample evaluation of MedSnap in Vientiane, Lao PDR (Laos) with the Medicine Quality Research Group at the Laos-Oxford-Mahosot-Wellcome Trust Research Unit (LOMWRU) of Mahosot Hospital.

Two MedSnap technicians visited the LOMWRU for three days. LOMWRU provided samples of azithromycin and ciprofloxacin directly obtained from trusted manufacturers and/or distributors in Laos, Cambodia, and Thailand. These *trusted* samples were assumed to be authentic for the purposes of this evaluation. LOMWRU also provided laboratory-tested samples of authentic and falsified artesunate and artemether-lumefantrine (Coartem). Field samples of ciprofloxacin and azithromycin were obtained in a convenience sample from local pharmacies in Vientiane City. All materials were organized and labeled by LOMWRU staff before being tested with the MedSnap technicians in a laboratory setting.

Trusted and authentic samples of all medicines were imaged with MedSnap DA and data was securely transferred to cloud servers for model building by technicians in the United States. A statistical model was created for each sample. Monte Carlo simulation methods were used to remove redundant models that were statistically indistinguishable from those retained. Retained models were securely transmitted to the remote MedSnap VR applications.

After trusted and authentic models were loaded, MedSnap VR was used to test each sample twice. In the case of a discrepancy between snap results, a third snap was performed and the majority result was counted.

Each VR snap was of a sample of one or more tablets, all with the same appearance. For brevity, we describe the process for tablet processing here. Capsule processing is similar with additional considerations for capsule caps, bodies, wrapped imprints, etc.

Tablets are placed on the Snap Surface, same-side-up according to the application prompt and imaged, then, if necessary, moved randomly and imaged again. Once enough images of the tablet-side are acquired (9 for MedSnap VR), the tablets are flipped and the same process is repeated.

With each snap, the applications first check for acceptable ambient lighting. Once the image is rectified and color-corrected, the application segments the image and extracts measurements of tablet features in five categories: shape, size, color, texture, and imprint. Each category is comprised of multiple features and in total, 25 features are computed per sample per tablet side. Each feature is averaged across the 9 images per tablet side and these average features are tested against their respective distributions in authentic models under the null hypothesis of no difference. If any test fails, the sample fails and is reported as not matching any authentic models. Multiple statistical tests are accommodated by the standard Holm-Bonferroni method^4^.

All samples of authentic and trusted medicines were tested with MedSnap VR. Eleven samples of known falsified artesunate of packaging types N°1,2,6,9,10,14 ^5^ and 14 samples of known falsified Coartem were also tested. In addition, 13 field samples of azithromycin and 8 field samples of ciprofloxacin were also tested.

## Results

One sample each of two visually distinct forms of authentic artesunate, 11 samples over 4 forms of trusted azithromycin, 8 samples over 6 forms of trusted ciprofloxacin, and 1 sample each of 2 forms of authentic artemether-lumefantrine (Coartem^®^ Novartis Pharmaceuticals Corporation, Suffern, New York, USA) were imaged with MedSnap DA and modelled.

Of the 48 samples tested, first-snap sensitivity was 100%, and first-snap specificity was 96%. One sample of trusted azithromycin 250mg returned a discrepant result between the two VR snaps. It was tested a third time and passed the trusted model yielding 2/3 pass. This discrepancy-resolved result was correct. Overall, sensitivity was 100% and discrepancy-resolved specificity was 100% (Table 1).

**Table 1.** MedSnap VR Results.

|  | Samples Pass | Samples Fail |  |
| --- | --- | --- | --- |
| Authentic/Trusted | **23** | **0** | **Specificity 100%** |
| Artesunate | 2 | 0 |  |
| Artemether-lumefantrine | 2 | 0 |  |
| Azithromycin | 11 | 0 |  |
| Ciprofloxacin | 8 | 0 |  |
| Falsified | **0** | **25** | **Sensitivity 100%** |
| Artesunate | 0 | 11 |  |
| Artemether-lumefantrine | 0 | 14 |  |

In addition, all 21 field samples of azithromycin and ciprofloxacin purchased from local pharmacies in Vientiane passed trusted models. Chemical confirmation of the field samples as authentic was not available, so the VR results from the field samples are not included in Table 1.

Analysis of visual relationships between authentic medications showed that within dose-form appearances, authentic samples were statistically indistinguishable.

**Figure 3.**
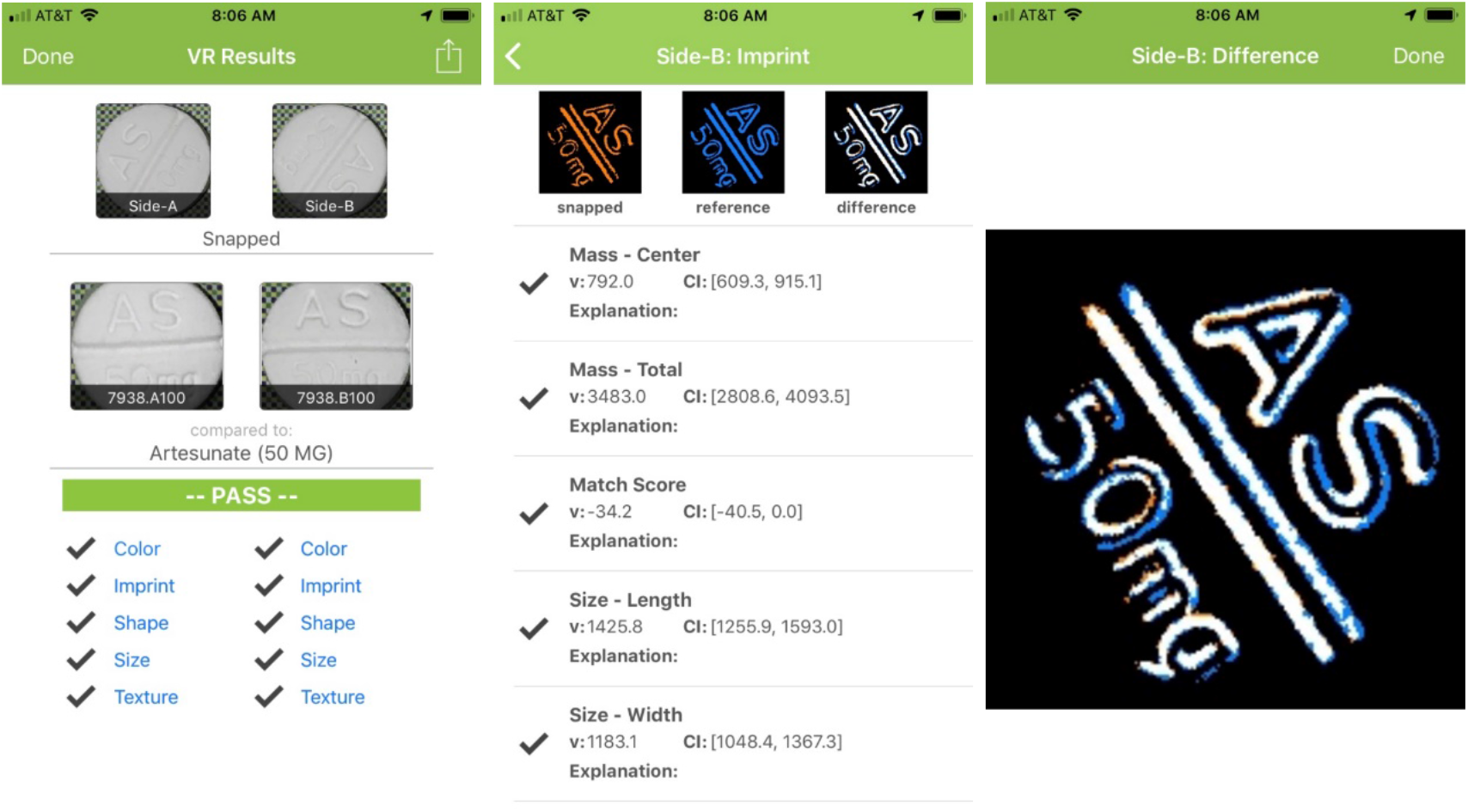
MedSnap VR allows the user to drill down into each imaging check performed. In the case above where the sample passed, the imprint composite of the sample and authentic model are being compared with an overlay (far right). Areas of exact match are in white, the sample is orange, and the reference is blue. This result is consistent with an exact imprint match.

**Figure 4.**
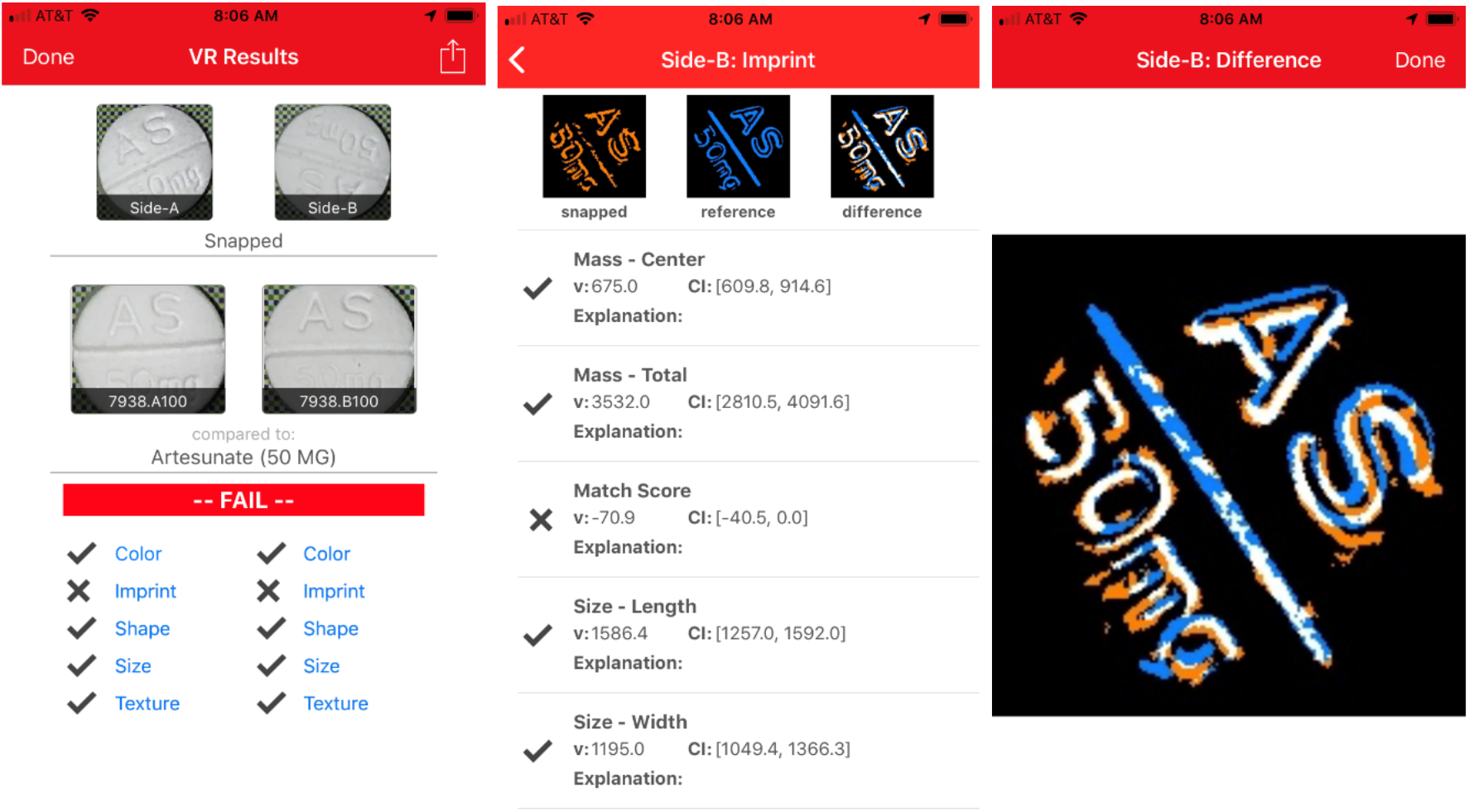
The above example shows a sample failing both A and B side imprint. Note that there is much less white in the overlay on the right, as the two composite imprints do not align. This can be due to materials used in manufacture, differences in dies, or how the tablet stamping machine was configured pre-production.

## Discussion

Modern medication manufacturing requires high-volume production in order to keep unit costs low. Mass production techniques for both tablets and capsules produce medicines with visual evidence of their authenticity, or lack thereof. Accordingly, the WHO recommends visual inspection as the first step to identifying potential counterfeit medicines^6^.

MedSnap uses computer vision, a consumer smartphone camera and a vinyl Snap Surface to model 25 visual features of solid oral medications. In this 3-day project in Lao PDR, authentic and trusted reference models were created, deployed, and used to test the local supply of two key medications (azithromycin and ciprofloxacin) along with archived samples of authentic and falsified artemether-lumefantrine and artesunate. MedSnap had a sensitivity of 100% and a specificity of 100% across 48 samples.

**Figure 5.**
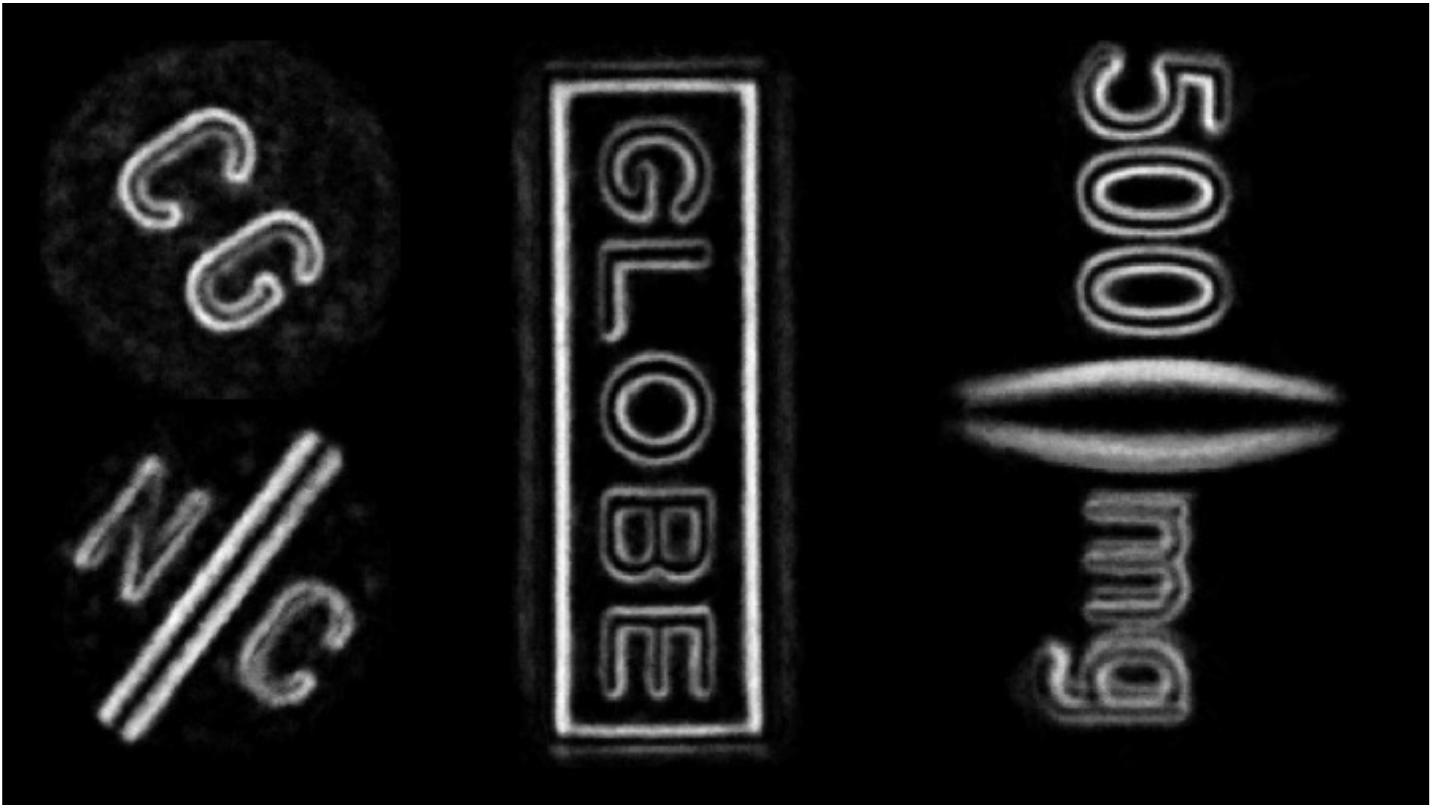
High resolution imprint composites are created during the modeling process to provide extremely accurate assessment of tablet or capsule imprint authenticity. Changes to tablet ingredients can affect how imprints are visualized due to changes in grain, reflectivity, or sharpness. From L->R, Coartem, Eurocapro, Cifloxin

MedSnap uses high intensity visual light from the smartphone flash and obtains precise color, size, and position measurements calibrated by the background Snap Surface. For example, tablets and capsules are measured in several dimensions to within +/− 0.1mm, over 250,000 shades of color can be captured, and highly detailed imprint composites are created and compared. Snap Surfaces can be cleaned with alcohol and are extremely durable.

Advantages of the MedSnap approach include handheld portability, ease of use for sample testing, and high precision measurement. In addition, central collection of detailed visual data on both authentic and falsified medications allows for prevalence analysis for public health and law enforcement.

Authentic samples can be imaged centrally by MedSnap staff or remotely by trained personnel. Authentic models are maintained in a secure, central library, and models created for one project can be selectively reused in others.

A typical project with a health system, regional healthcare provider, or government (sponsor) proceeds as follows:

- Engagement with the sponsor to determine
  - Medications to be surveilled and/or tested with the MedSnap VR application
  - Smartphone models to be supported
  - Venues in which to deploy the application e.g. clinics, pharmacies, regulatory agencies, law enforcement
  - Number of anticipated users and their training requirements
  - Language
  - Any customized data elements required
- Samples of authentic medications selected, optimally sent directly from the manufacturer or distributor, are stored in a central location chosen by the sponsor, or to MedSnap
- Authentic medications are imaged by select personnel using MedSnap DA
- All authentic model data and images are reviewed by MedSnap QA personnel
- Authentic models are built
- Authentic models are deployed to MedSnap VR users within the project
- Project specific support and training personnel interact with end users to support appropriate use
- Monthly reports of use by location, user, medication, result (authentic, falsified)
- Optional quarterly reports detailing the prevalence of distinct falsified variants

MedSnap can be used for ongoing screening surveillance and targeted investigations. Administrators of MedSnap projects can decide what medicines to include and where to deploy MedSnap VR.

The system is appropriate for use by clinic or pharmacy personnel when medication supplies need to be authenticated or when medicines need to be checked on suspected treatment failure. In parts of the world with high prevalence of falsified medications, encouraging patients to bring their medications to clinic or to the pharmacy for authentication could become another vital sign to ensure their recovery and improve the overall efficiency of care.

### Limitations of the study

We assumed that samples of azithromycin and ciprofloxacin, acquired from trusted manufacturers and distributors, were authentic. Laboratory chemical testing was not performed on these samples. Laboratory testing was performed on curated authentic and falsified samples of artesunate and Coartem. Only four medicines, with five active pharmaceutical ingredients, and only tablets were included. The falsified and authentic samples of artemether-lumefantrine and artesunate that were tested were well beyond expiry dates (at least 3 years past expiry) which may have altered the physical appearance of the tablets and distorted the results.

## Conclusion

MedSnap is an accurate, scalable application requiring only a handheld, widely available consumer smartphone and a durable, vinyl background for sample testing. It can be used in developing and developed markets to screen solid oral medications along the supply chain, including at the point of dispensing in the pharmacy or clinic. Use of MedSnap in combination with other chemical testing devices to evaluate API may increase confidence in the MedSnap authentic medication library. The ability for local or regional users to specify likely authentic medications as such and rapidly deploy models for surveillance is an advantage for the system.

## Data Availability

The data that support the findings of this study are available from the corresponding author, upon reasonable request.

## Acknowledgements

This project was completed through a partnership between Indicator Sciences (Seattle, WA, USA, www.medsnap.com), Becton Dickinson (Franklin Lakes, NJ, USA, www.bd.com), the Foundation for Innovative New Diagnostics (FIND, Geneva, Switzerland, www.finddx.org), and the Lao-Oxford-Mahosot Hospital Wellcome Trust Research Unit (LOMWRU, Vientiane, Laos).

The overall project, coordinated by FIND, is assessing deployment models for field-based diagnostics to reduce antibiotic resistance caused by substandard and falsified medicines.

Special thanks to Professor Paul Newton, Dr. Céline Caillet, Vayouly Vidhamaly, Kem Boutsamay, the staff at LOMWRU, and the Director and staff of the Microbiology Laboratory, Mahosot Hospital, for their assistance in completing this assessment.

This project is supported by the Fleming Fund, a UK aid programme from the UK’s Department of Health and Social Care (www.flemingfund.org)

Correspondence regarding this work should be sent to Dr. Patrick Hymel

## Conflict of Interest Statement

The authors are employed full time with Indicator Sciences. Drs. Hymel and Brossette are the owners of Indicator Sciences.

Indicator Sciences received only travel and hardware expense reimbursement for this work.

## Appendix

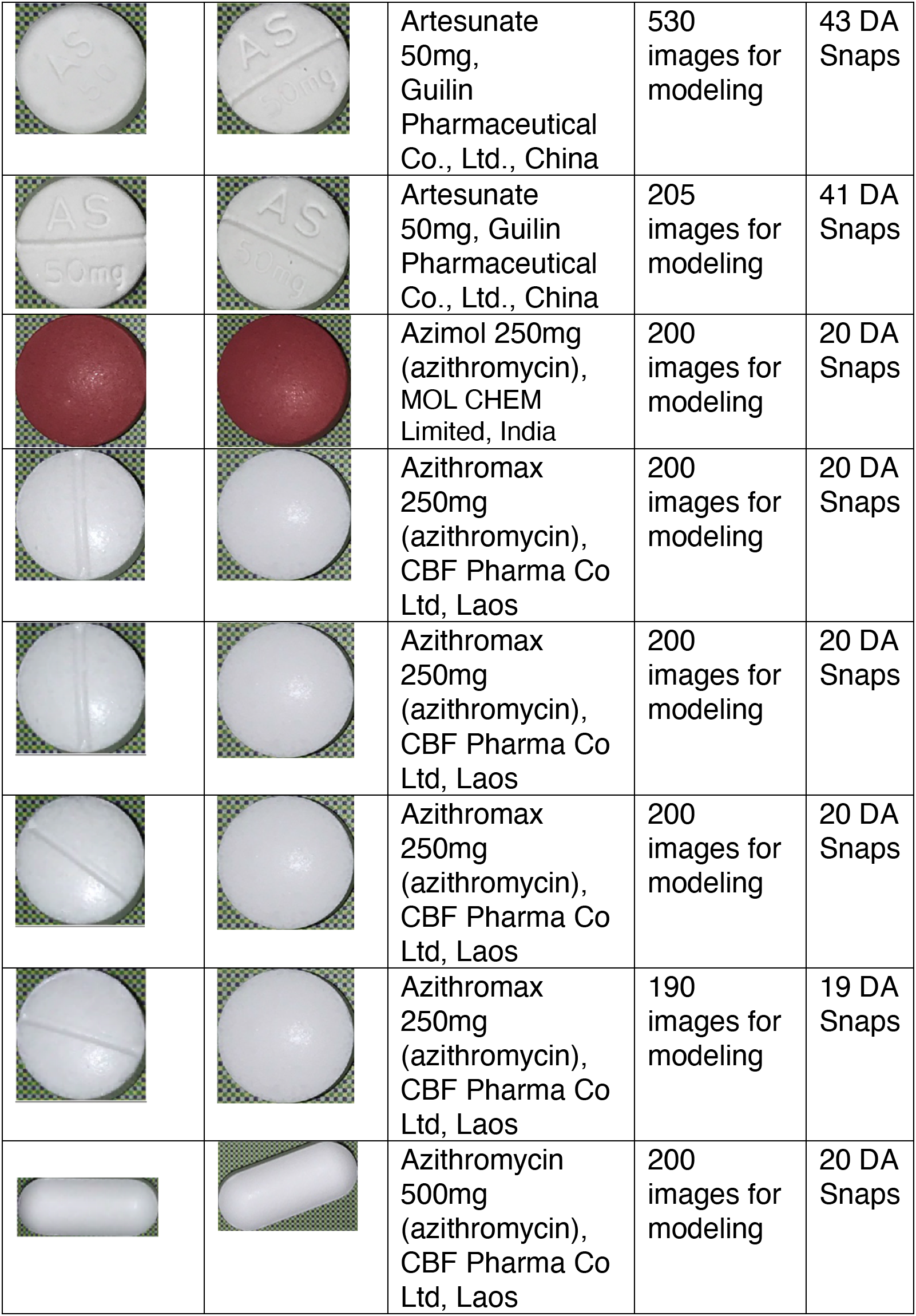
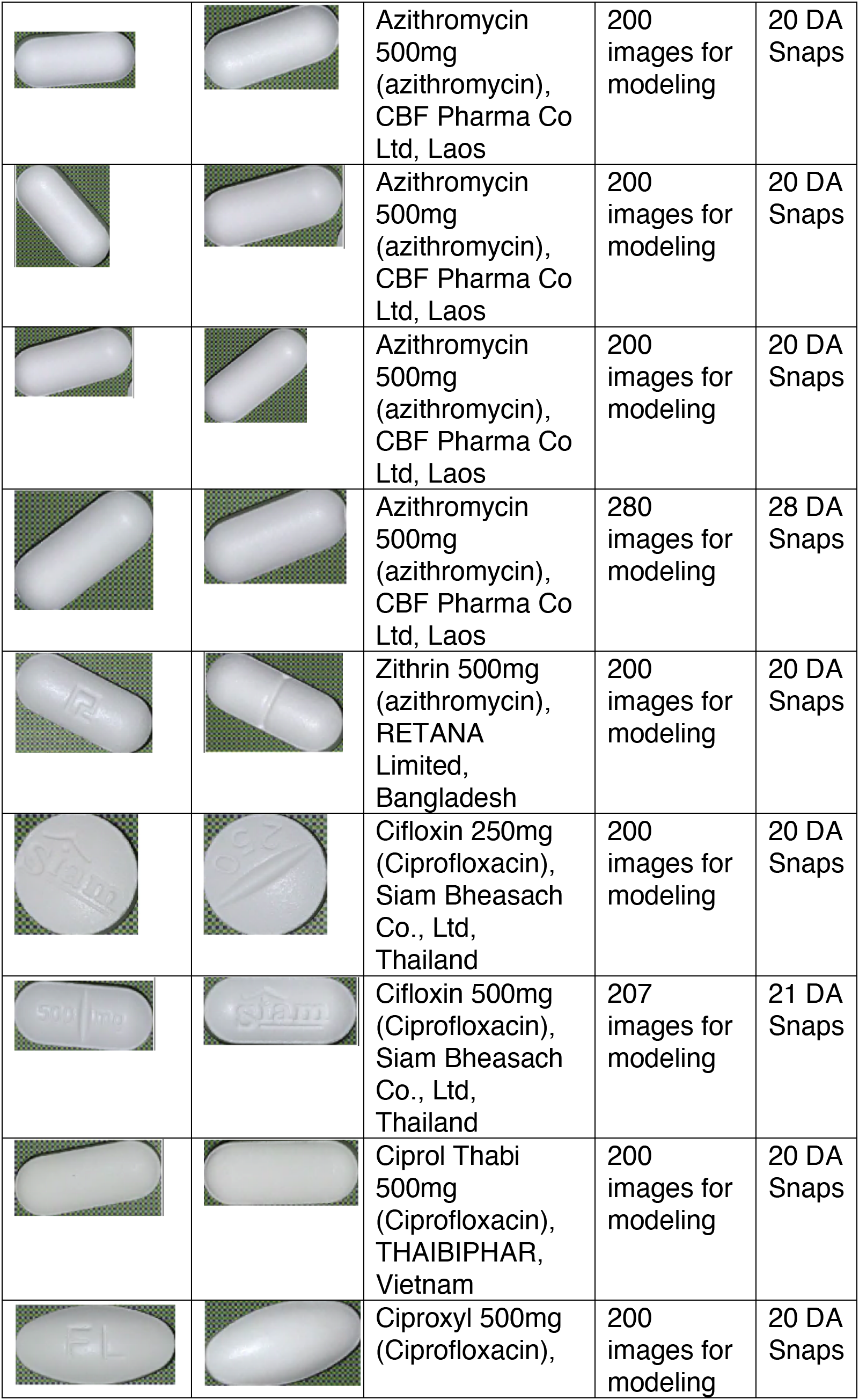
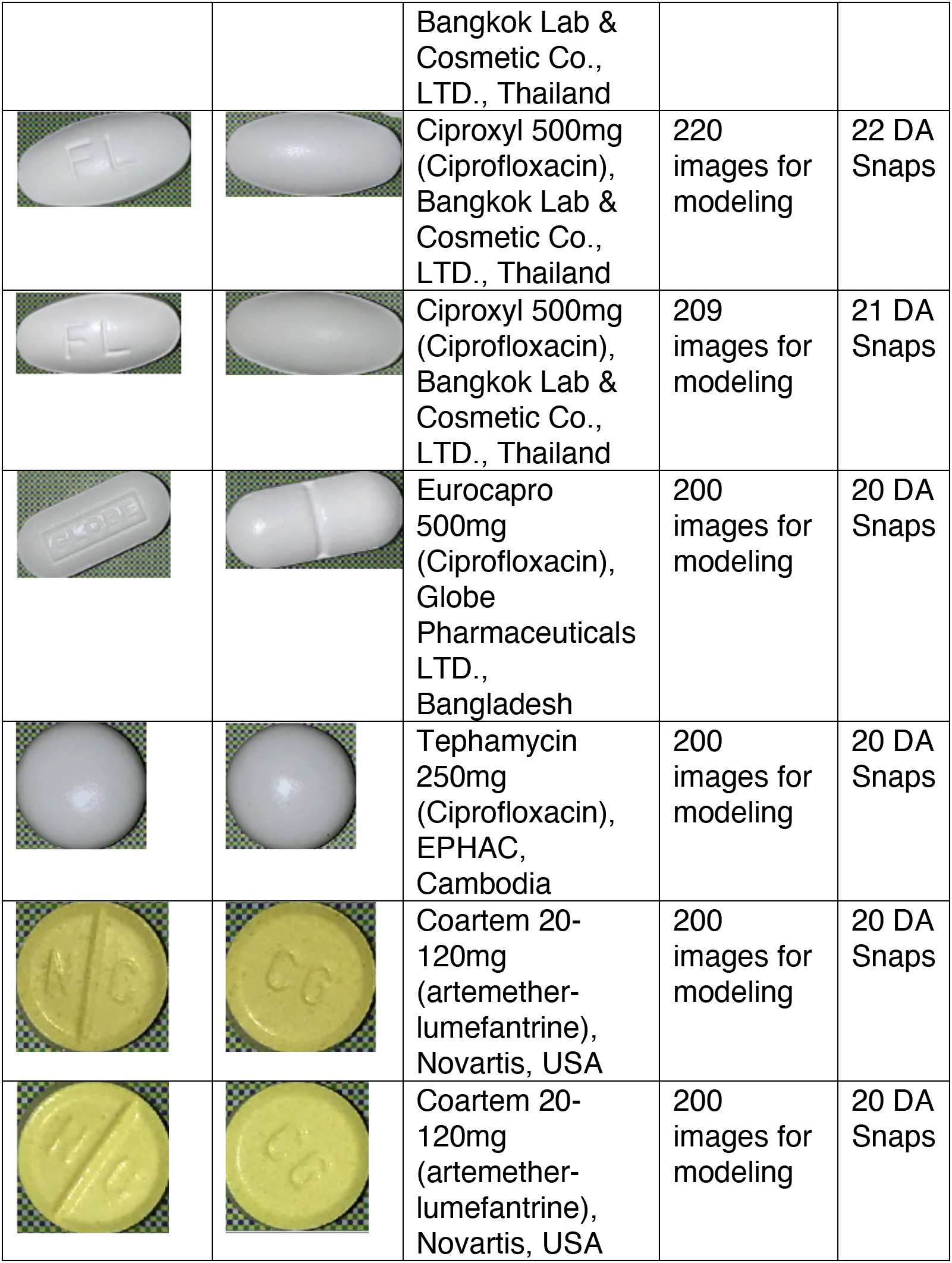
Authentic and trusted samples imaged with the MedSnap DA for authentic model building.

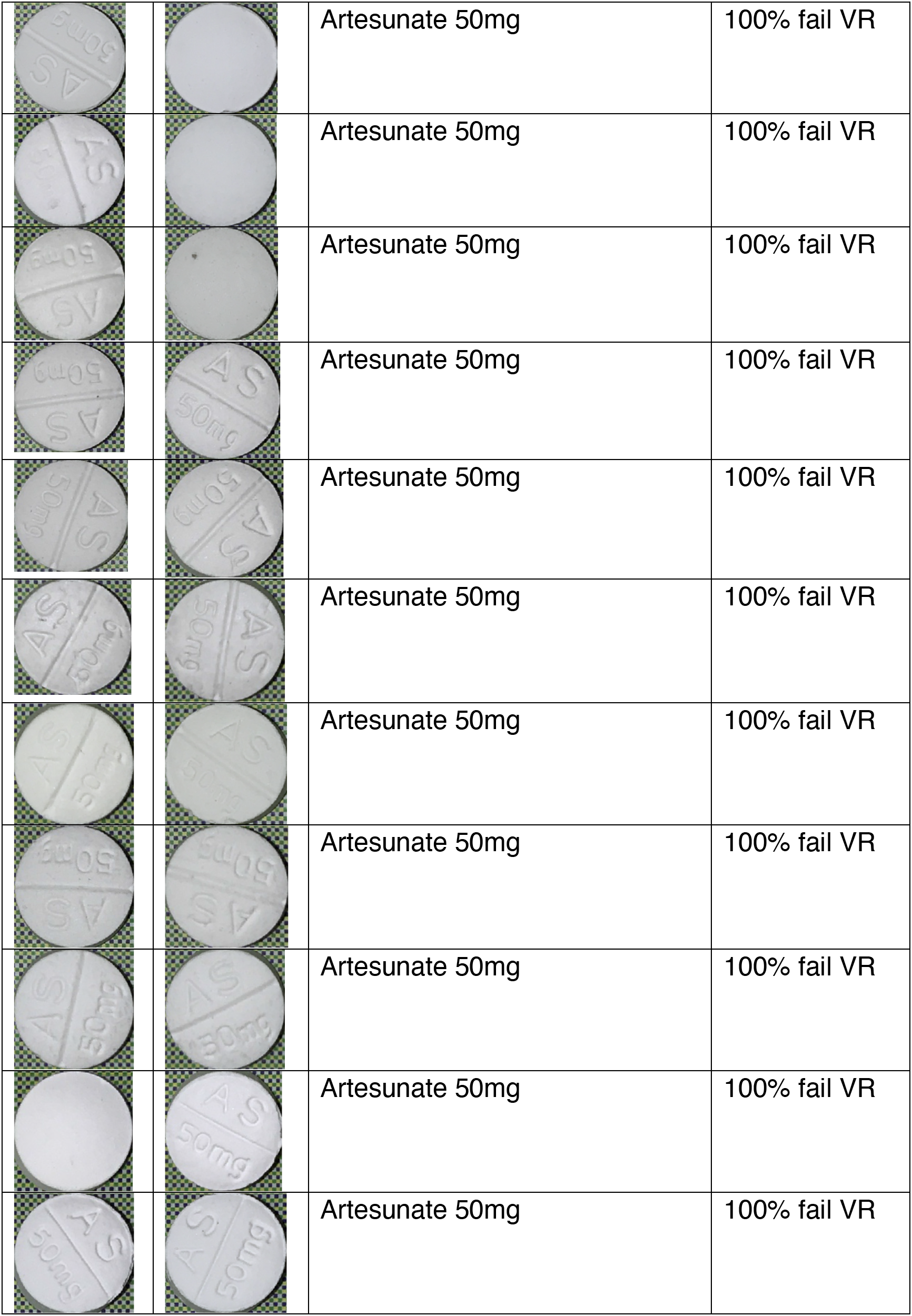
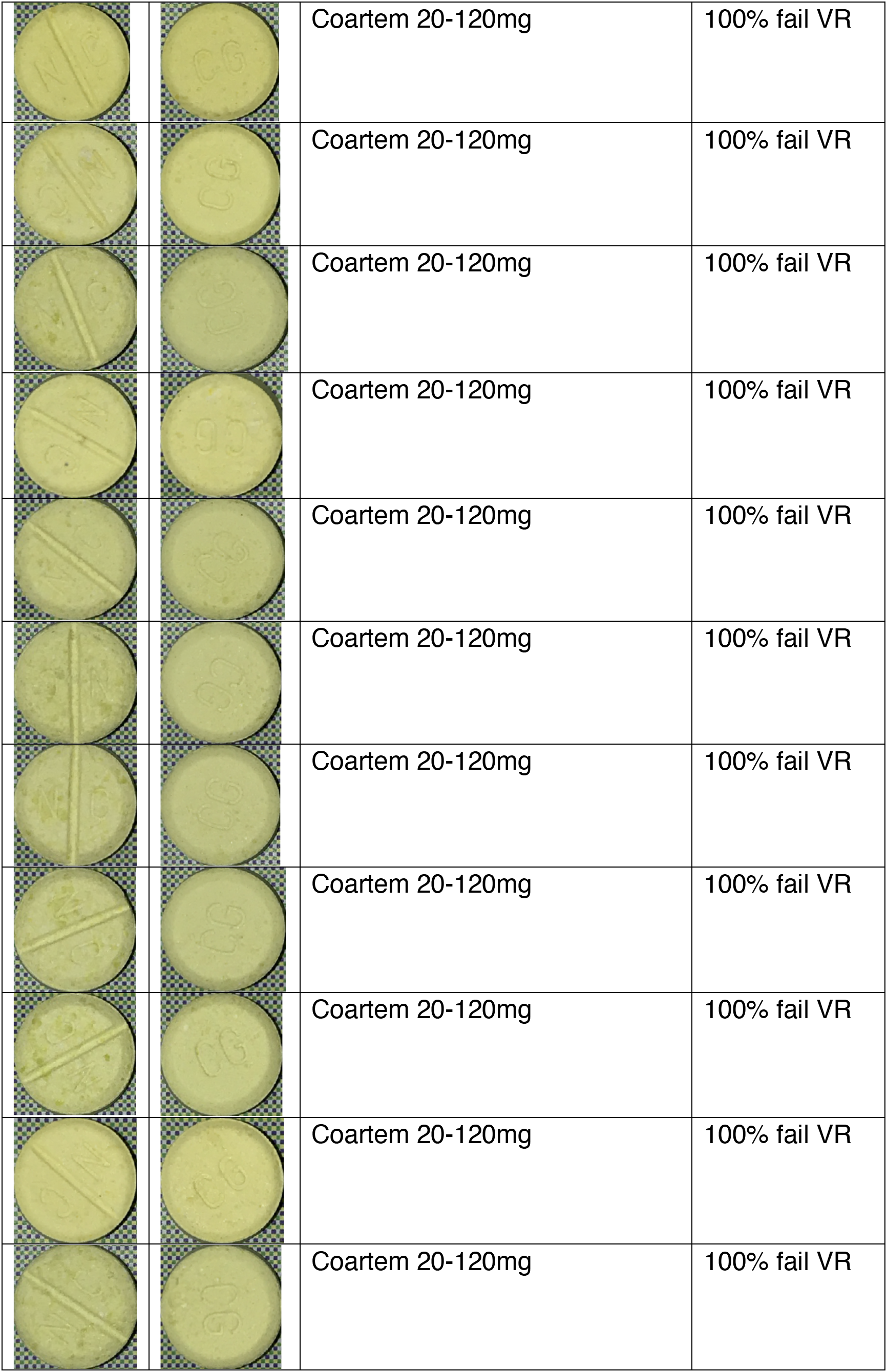
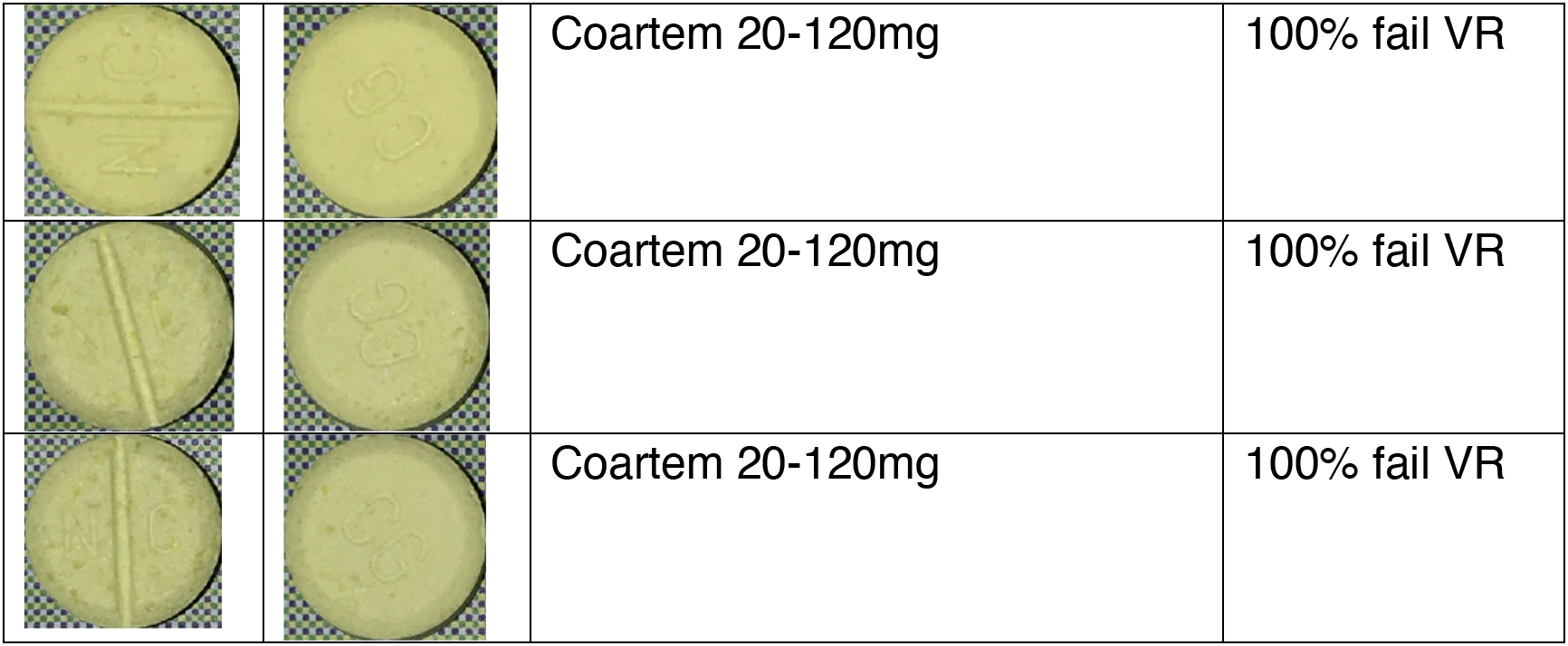
Falsified Tablets Tested With MedSnap VR.

## References

1 WHO Global Surveillance and Monitoring System for substandard and falsified medical products. Geneva: World Health Organization; 2017. Licence: CC BY-NC-SA 3.0 IGO.

2 “Medical Product Alert N°3/2020 Falsified medical products, including in vitro diagnostics, that claim to prevent, detect, treat or cure COVID-19.” WHO, https://www.who.int/docs/default-source/essential-medicines/drug-alerts20/no3-2020-falsified-mp-forcovid-en.pdf?sfvrsn=cd86600116. Accessed 5 May 2020.

3 Vickers S, et al. Field detection devices for screening the quality of medicines: a systematic review. BMJ Glob Health 2018;3:e000725. doi:10.1136/bmjgh-2018-000725

4 Holm, S. (1979). “A simple sequentially rejective multiple test procedure”. Scandinavian Journal of Statistics. 6 (2): 65–70. JSTOR 4615733.

5 Newton PN, Fernández FM, Plançon A, Mildenhall DC, Green MD, et al. (2008) A collaborative epidemiological investigation into the criminal fake artesunate trade in South East Asia. PLoS Med 5(2): e32. doi:10.1371/journal.pmed.0050032

6 WHO Guidelines for the Development of Measures to Combat Counterfeit Drugs. Geneva. World Health Organization; 1999.

